# Obfuscation via pitch-shifting for balancing privacy and diagnostic utility in voice-based cognitive assessment

**DOI:** 10.1101/2024.11.25.24317900

**Authors:** Meysam Ahangaran, Nauman Dawalatabad, Cody Karjadi, James Glass, Rhoda Au, Vijaya B. Kolachalama

## Abstract

**Introduction:** Digital voice analysis is gaining traction as a tool to differentiate cognitively normal from impaired individuals. However, voice data poses privacy risks due to the potential identification of speakers by automated systems.

**Methods:** We developed a framework that uses weighted linear interpolation of privacy and utility metrics to balance speaker obfuscation and cognitive integrity in cognitive assessments. This framework applies pitch-shifting for speaker obfuscation while preserving cognitive speech features. We tested it on digital voice recordings from the Framingham Heart Study (N=128) and Dementia Bank Delaware corpus (N=85), both containing responses to neuropsychological tests.

**Results:** The tool effectively obfuscated speaker identity while maintaining cognitive feature integrity, achieving an accuracy of 0.6465 in classifying individuals with normal cognition, mild cognitive impairment, and dementia in the FHS cohort.

**Discussion:** Our approach enables the development of digital markers for dementia assessment while protecting sensitive personal information, offering a scalable solution for privacy-preserving voice-based diagnostics.

## Background

Voice recordings contain valuable information that can indicate an individual’s cognitive health,^1^ offering a non-invasive and efficient method for assessment. Research has shown that digital voice markers can detect early signs of cognitive decline by analyzing features such as speech rate, articulation, pitch variation, and pauses, which may signal cognitive impairments when deviating from normative patterns.^2^ These markers, combined with additional derived measures, have been effective in distinguishing between normal cognition (NC), mild cognitive impairment (MCI), and dementia (DE). Advancements in data-driven frameworks, particularly machine learning, have further enhanced the utility of voice markers by enabling the identification of complex patterns associated with cognitive states.^3–11^ Machine learning models can analyze large datasets of voice samples to detect subtle changes, providing an objective and scalable approach to cognitive assessment.^12–14^ This technology holds potential for early screening in clinical settings as well as remote monitoring, which could be especially valuable in resource-limited environments. Consequently, voice-based diagnostics are emerging as a promising complement to traditional assessments like neuropsychological tests and neuroimaging.

The use of voice data as an early diagnostic tool for neurodegenerative diseases like Alzheimer’s is increasingly relevant in today’s aging population, where early detection of cognitive decline can improve patient outcomes through timely interventions.^15^ However, voice data introduces privacy challenges due to the personally identifiable information (PII) embedded in recordings, such as gender, accent, and emotional state, as well as subtler speech characteristics that can re-identify individuals. These risks are amplified when voice data is processed by automated systems,^16^ raising concerns about re-identification and data misuse. This has led to a demand for privacy-preserving techniques that protect speaker identity while maintaining the critical speech features necessary for accurate cognitive health assessments. Traditional anonymization techniques, such as voice scrambling or noise addition, often degrade acoustic features vital for clinical analysis, making it challenging to balance privacy with diagnostic utility. Current privacy-preserving methods typically focus on either masking speaker identity or preserving speech intelligibility, but they often overlook the need to retain cognitive markers crucial for dementia diagnosis. For example, features like voice pitch, prosody, and articulation speed, while important for detecting cognitive decline, can also reveal speaker identity. Consequently, many existing methods either sacrifice diagnostic accuracy for privacy or inadequately protect identity. The challenge lies in developing an approach that obfuscates speaker identity without compromising the cognitive features necessary for assessing cognitive health.

In this study, we developed a framework that utilizes pitch-shifting techniques for voice obfuscation, aiming to reduce the risk of speaker identification while preserving the integrity of cognitive features relevant for dementia assessment (**Fig. 1**). This approach was applied to voice data from the Framingham Heart Study and the Dementia Bank Delaware corpus, both containing spoken responses to neuropsychological tests. We leveraged six classification algorithms to assess the preservation of cognitive speech features following obfuscation. Our approach emphasizes the need to balance privacy and cognitive utility in voice recordings, particularly for diagnostic applications. We employed weighted linear interpolation to calibrate the trade-off between speaker privacy and cognitive feature utility, ensuring that obfuscation did not significantly compromise the diagnostic value of speech features. This approach addresses the challenge of using voice data for dementia assessments while mitigating privacy risks.

**Figure 1.**
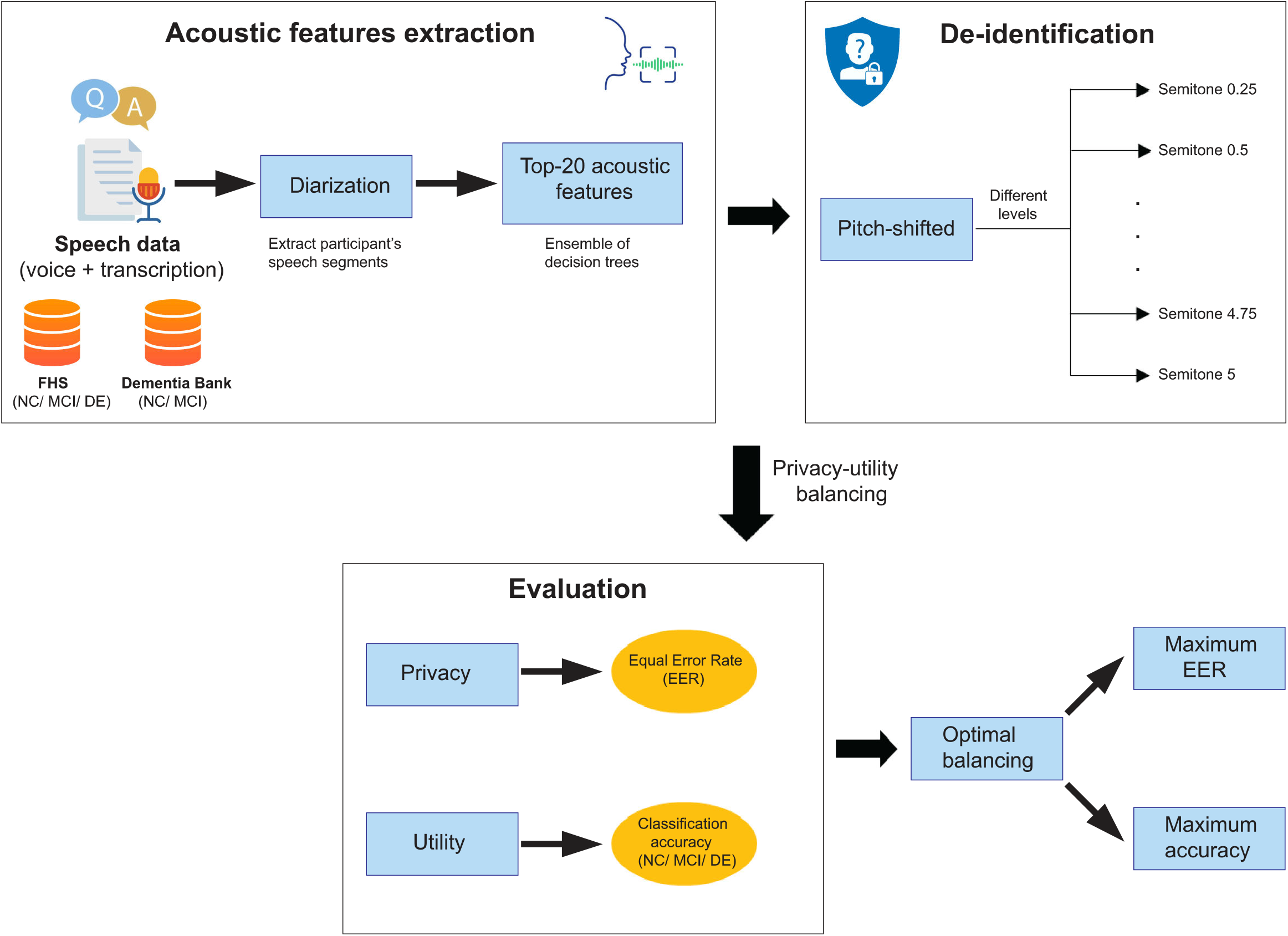
Privacy-utility balancing framework applied to digital voice recordings for cognitive assessments. This figure presents an overview of the proposed speaker obfuscation strategy using a pitch-shifting method aimed at obscuring speaker identity while preserving cognitive features critical for dementia diagnosis. The framework was tested on neuropsychological voice recordings from the Framingham Heart Study and the Dementia Bank Delaware corpus. Six classification algorithms were employed to assess cognitive integrity across three cognitive groups: normal cognition (NC), mild cognitive impairment (MCI), and dementia (DE).

## Methods

### Study population

We applied obtained voice recordings from the Framingham Heart Study (FHS) and the Dementia Bank Delaware corpus.^8, 9, 17–19^ The FHS dataset included 128 speech samples from participants with either NC, MCI, or DE. The Dementia Bank dataset included 85 speech samples, with either NC or MCI participants. FHS is a community-based longitudinal observational study initiated in 1948, which has provided significant insights into the epidemiology of cardiovascular disease and its risk factors across multiple generations.^19^ The FHS dataset encompasses a variety of neuropsychological tests that assess multiple cognitive domains, including memory, attention, executive function, language, reasoning, visuoperceptual skills, and premorbid intelligence. The cognitive status of participants, classified as NC, MCI, or DE, was determined over time by the FHS dementia diagnosis review panel. For each participant, cognitive status was assigned based on the diagnosis date closest to the recording date, either on or before the recording date, or within 180 days thereafter.

The FHS speech files, originally in WAV format (74.36 ± 26.62 minutes, sampling rate 22,050 Hz), and the Dementia Bank data, originally in MP3 format (10.81 ± 4.82 minutes), were converted to WAV format with sampling rate of 22,050 Hz to ensure consistency in processing. Both datasets included recordings of interactions between clinicians and participants, encompassing both questions and responses (**Table 1**). Speech diarization was applied to isolate participant speech by removing clinician interactions, ensuring the analysis focused on vocal features relevant to cognitive impairment.

**Table 1.** Study population. Demographic and cognitive status characteristics of the Framingham Heart Study (FHS) and Dementia Bank datasets. The table provides information on the number of participants (N), cognitive status classification (Normal Cognition [NC], Mild Cognitive Impairment [MCI], Dementia [DE]), percentage of male participants, average age with standard deviation (mean ± std), and average speech duration in minutes (mean ± std) for both datasets.

| Dataset | Cognitive status | Male percentage (%) | Age (mean±std) | Speech duration (mean±std minutes) |
| --- | --- | --- | --- | --- |
| **FHS**<br>**(N = 128)** | NC (N=40) | Male = 60 (47%) | 82.84 ± 8.22 | 74.36 ± 26.62 |
| | MCI (N=10) | NC = 25 (19%) | NC = 80.37 ± 10.46 | NC = 75.05 ± 25.3 |
| | DE (N=78) | MCI = 6 (5%) | MCI = 80.3 ± 7.63 | MCI = 57.83 ± 24.08 |
| | | DE = 29 (23%) | DE = 84.43 ± 6.39 | DE = 76.13 ± 26.87 |
| **Dementia**<br>**bank**<br>**(N = 85)** | NC (N=34) | Male = 27 (32%) | 71.36 ± 7.38 | 10.81 ± 4.82 |
| | MCI (N=51) | NC = 5 (6%) | NC = 68.23 ± 5.82 | NC = 12.1 ± 5.7 |
| | | MCI = 22 (26%) | MCI = 73.45 ± 7.57 | MCI = 9.94 ± 3.91 |

### Derivation of acoustic features from speech recordings

We extracted acoustic features from speech recordings using the Python *librosa* package at a 22,050 Hz sampling rate for high-fidelity capture.^20^ A total of 12 distinct sets of vocal features were derived, including statistical measures such as minimum, maximum, mean, standard deviation, and median, resulting in 481 features. The extracted features included parameters like amplitude, root mean square, spectral coefficients, bandwidth, centroid, flatness, roll-off frequency, zero-crossing rate, tempo, Chroma Energy Normalized (CENS), and Mel-Frequency Cepstral Coefficients (MFCC) with delta features. The MFCC delta features were calculated using Savitzky-Golay filtering to capture temporal derivatives.^21^

### Balancing privacy and cognitive integrity in speech de-identification

Our framework sought to balance speaker privacy and cognitive data preservation by dividing the feature set into two subsets: cognitive features crucial for dementia diagnosis and obfuscation features related to speaker identity. We enhanced privacy by modifying the obfuscation features while maintaining the integrity of cognitive features. To achieve this balance, we used two performance metrics: privacy degradation, measured by the Equal Error Rate (EER) of an Automatic Speaker Verification (ASV) system,^22^ and cognitive utility, measured by the accuracy (ACC) of a machine learning model that differentiated cognitive states. We applied weighted linear interpolation to combine EER and ACC, allowing us to explore different obfuscation levels and find the optimal trade-off between privacy and cognitive utility. Specifically, we quantified the overall performance of obfuscation (ρ) by incorporating the privacy level (α), within the range 0 ≤ *α* ≤ 1, using the formula:

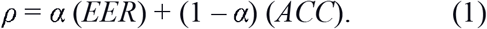

The value of *ρ* ranges from 0 to 1 and setting *α* to 0.5 gave equal importance to privacy and cognitive utility.

Using a pitch-shifting method, we altered speech data at varying levels from 0 to 5 semitones. The pitch shifts were normalized on a scale from 0.05 to 1, creating 20 different configurations to assess the impact of these shifts on both privacy (speaker obfuscation) and utility (cognitive classification). At each pitch-shifting level, we calculated the classification accuracy across six different machine learning algorithms (Random Forest, Support Vector Machine (SVM), k-Nearest Neighbors (kNN), Multi-Layer Perceptron (MLP), AdaBoost, and Gaussian Naive Bayes) to assess how changes in acoustic features affected cognitive differentiation. To quantify privacy, we used the EER from an ASV system to compare original and obfuscated files. We also calculated the Area Under the Curve (AUC) for binary classification in speaker recognition, with lower AUC and higher EER values indicating more effective obfuscation.

## Results

### Acoustic features for cognitive assessment

The FHS dataset had a longer average speech duration (74.36 ± 26.62 minutes) compared to the Dementia Bank dataset (10.81 ± 4.82 minutes). Longer samples provide a broader range of acoustic features and more detailed vocal characteristics. After extracting acoustic features, we aimed to identify the most significant vocal features relevant for dementia diagnosis. Using supervised learning, we labeled each participant’s speech data based on their cognitive status. We applied random forest regression with an ensemble of 100 decision trees, to evaluate the contribution of each feature to predictive performance. The resulting importance scores were normalized between 0 and 1, and then ranked.

We selected the top 20 acoustic features with the highest importance scores from both datasets for further analysis (**Supplementary Data 1** and **2**). In the FHS dataset, the key features included statistical measures of MFCC, MFCC delta, zero-crossing rate, CENS, and spectral bandwidth (**Fig. 2a**). Similarly, in the Dementia Bank dataset, the most important features were statistical measures of MFCC, MFCC delta, and CENS (**Fig. 2.b**). We evaluated how the number of selected acoustic features influenced classification accuracy. We iteratively selected the top-k features, ranging from k=1 to k=481, and applied six classification methods: Random Forest, Support Vector Machine (SVM), k-Nearest Neighbors (kNN), Multi-Layer Perceptron (MLP), AdaBoost, and Gaussian Naive Bayes, using 10-fold cross-validation (**Supplementary Data 3** and **4**). The tasks included differentiating between NC, MCI, and DE in the FHS dataset, and NC and MCI in the Dementia Bank dataset.

**Figure 2.**
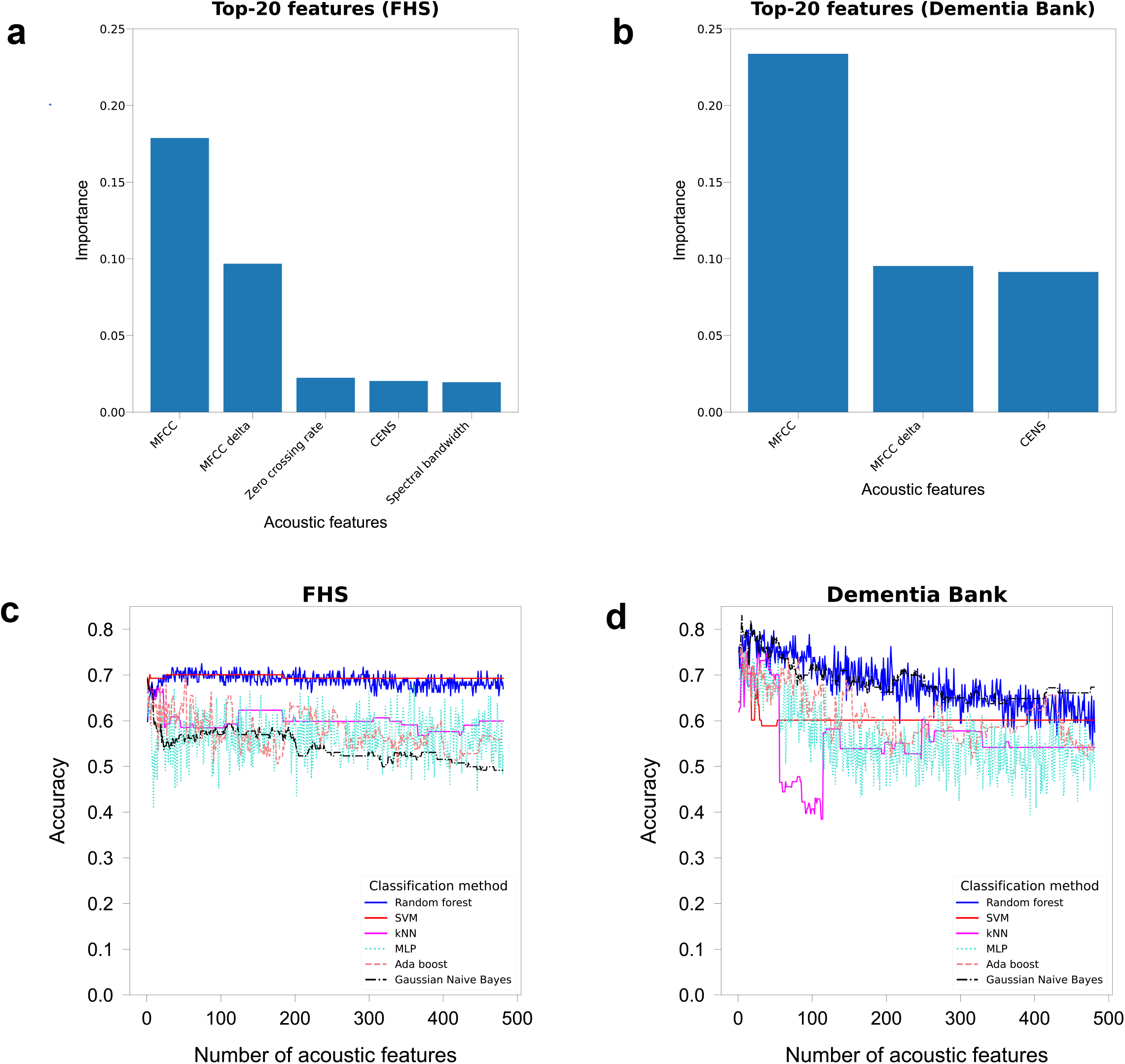
Selection and impact of key acoustic features. (a) Importance ranking of the top 20 acoustic features from the FHS dataset, determined using random forest regression. (b) Importance ranking of the top 20 acoustic features from the Dementia Bank dataset, determined using random forest regression. (c) Effect of the number of selected acoustic features on classification accuracy for NC, MCI, and DE across six classification algorithms in the FHS dataset. (d) Effect of the number of selected acoustic features on classification accuracy for NC and MCI across six classification algorithms in the Dementia Bank dataset.

In the FHS dataset, Random Forest and SVM reached an accuracy of approximately 0.7 after employing 20 features, with kNN showing a slight decrease in accuracy with further feature inclusion. MLP exhibited the most fluctuation across all feature sets (**Fig. 2c**). In the Dementia Bank dataset, all algorithms showed a decrease in accuracy as more features were included, with Random Forest and Gaussian Naive Bayes displaying minimal variation (**Fig. 2d**). We also performed classification on both the original and top-20 acoustic features. Across the six algorithms, using the top-20 features improved average classification accuracy by 0.0149 and 0.1523 in the FHS and Dementia Bank datasets, respectively (**Supplementary Table 1**). Additionally, an analysis with Random Forest yielded improved AUC scores for cognitive category differentiation, with increases of up to 0.2 for NC/MCI binary classification in the Dementia Bank dataset (**Fig. 3**). These results suggest that a reduced set of top-ranked features can enhance classification accuracy while maintaining computational efficiency.

**Figure 3.**
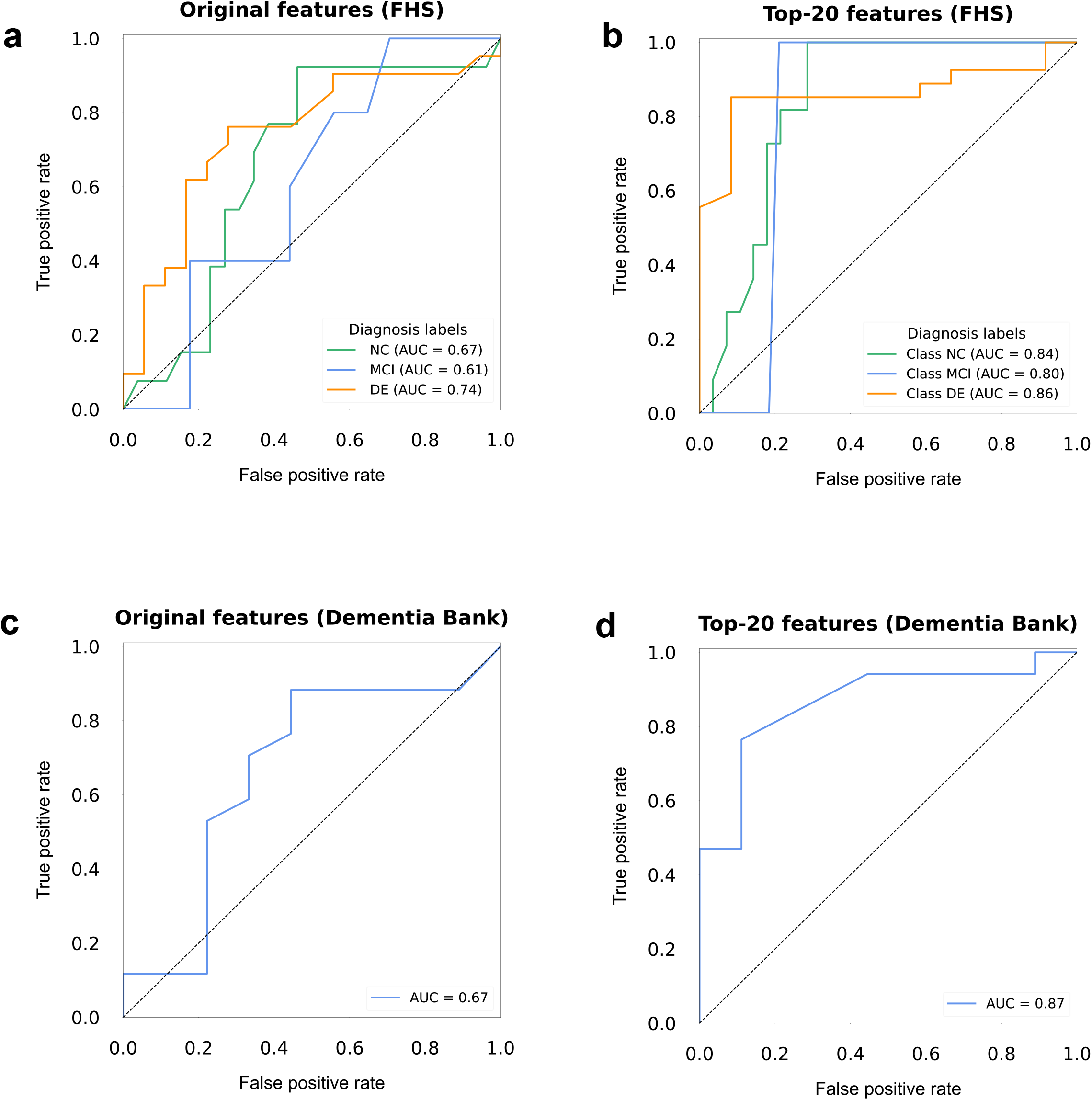
Classification performance using original and top-20 acoustic features. (a) ROC curves and AUC scores for NC, MCI, and DE classification in the FHS dataset using the original feature set and Random Forest classifier. (b) ROC curves and AUC scores for NC and MCI classification in the Dementia Bank dataset using the original feature set and Random Forest classifier. (c) ROC curves and AUC scores for NC, MCI, and DE classification in the FHS dataset using the top-20 features and Random Forest classifier. (d) ROC curves and AUC scores for NC and MCI classification in the Dementia Bank dataset using the top-20 features and Random Forest classifier.

### Evaluation of privacy-utility balancing in acoustic feature manipulation

Our results demonstrate how varying pitch-shifting levels affected both classification accuracy and privacy metrics (**Fig. 4**). Classification accuracy, EER scores, and AUC values provided insights into how varying pitch-shifting levels impacted the trade-off. **Figures 4a** and **4b** show the model’s performance across privacy levels α, ranging from 0.05 to 1. Optimal performance was achieved at privacy levels of 0.35 for the Framingham Heart Study (FHS) dataset (pitch-shifting level 0.4) and 0.3 for the Dementia Bank dataset (pitch-shifting level 0.85), as indicated by vertical orange dashed lines (**Supplementary Tables 2** and **3**). **Figures 4c** and **4d** illustrate the variation in model performance with increasing pitch-shifting levels, including metrics such as EER, classification accuracy, and AUC. The FHS dataset achieved a total performance score (ρ) of 0.5372 at a pitch-shifting level of 0.4 and privacy level *α* of 0.35 (**Supplementary Data 5**). In the Dementia Bank dataset, a pitch-shifting level of 0.85 resulted in a total performance score of 0.5218 with privacy level *α* of 0.3 (**Supplementary Data 6**). **Figures 4e** and **4f** provide heatmaps showing total performance *ρ* at different pitch-shifting levels, along with corresponding EER scores. The optimal pitch-shifting levels for both datasets were 0.4 (ρ = 0.5372, EER = 0.3342, ACC = 0.6465) for FHS and 0.85 (ρ = 0.5218, EER = 0.2908, ACC = 0.6208) for Dementia Bank. **Supplementary Figure 1** further illustrates the correlations between pitch-shifting levels and various privacy and utility metrics for the FHS and Dementia Bank datasets.

**Figure 4.**
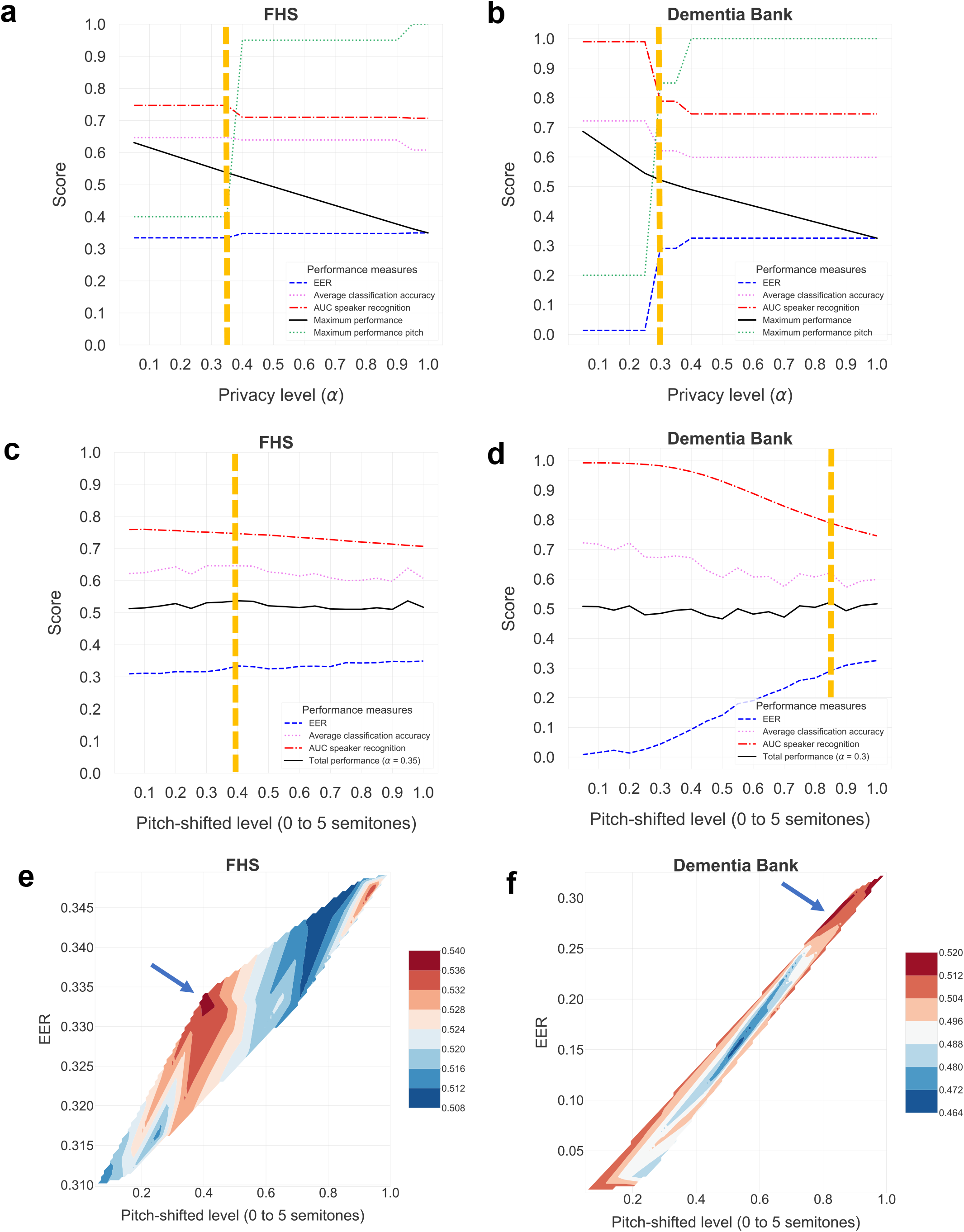
Privacy-utility analysis. (a) Model performance assessment across different privacy levels in the FHS dataset, showing that the optimal balance occurs at a privacy level of *α* = 0.35, corresponding to a pitch-shifting level of 0.4. (b) Model performance evaluation across varying privacy levels in the Dementia Bank dataset, with the optimal balance achieved at a privacy level of *α* = 0.3, corresponding to a pitch-shifting level of 0.85. (c) Impact of increasing pitch-shifting levels on model performance across different metrics in the FHS dataset. (d) Effect of increasing pitch-shifting levels on model performance across various metrics in the Dementia Bank dataset. (e) Contour heat map showing total performance *ρ* at different pitch-shifting levels in the FHS dataset. (f) Contour heat map illustrating total performance *ρ* at different pitch-shifting levels in the Dementia Bank dataset.

The results from this study provide insights into how pitch-shifting can balance privacy and cognitive utility in dementia diagnosis using voice data. Optimal pitch-shifting levels (0.4 for FHS and 0.85 for Dementia Bank) effectively preserved cognitive feature utility while achieving sufficient speaker obfuscation. As demonstrated by the total performance scores (ρ) and EER values, the trade-off between speaker privacy and diagnostic accuracy was managed successfully, allowing us to maintain diagnostic utility without compromising privacy.

## Discussion

Our findings demonstrate that varying levels of pitch-shifting can affect speaker privacy and the utility of acoustic features for assessment of cognitive status. The differences in optimal pitch-shifting levels between the FHS and Dementia Bank datasets are attributable to factors such as data size and feature sensitivity. The longer speech samples in the FHS dataset provided more robust performance in balancing privacy and utility, while the Dementia Bank dataset required more substantial pitch-shifting due to shorter duration and greater feature sensitivity. Overall, these findings underscore the importance of tailoring obfuscation techniques to the characteristics of the dataset being analyzed.

Despite these promising results, our study has a few limitations. First, pitch-shifting may alter acoustic features crucial for diagnosis, potentially compromising classification accuracy. Future research should explore more advanced obfuscation techniques that preserve diagnostic utility while ensuring robust privacy protection. Additionally, reliance on ASV systems for de-identification effectiveness may lead to overestimation of privacy protection, as these systems do not fully capture the nuances of altered speech. Incorporating human perception studies or adversarial models could provide further insights into the robustness of de-identification. Our study also focused on a limited set of acoustic features and classification algorithms. Future work should explore a broader range of features, including temporal and prosodic elements, and additional machine learning methods to enhance classification accuracy across diverse populations. Expanding the research to more diverse cohorts would strengthen the generalizability and clinical applicability of pitch-shifting techniques. Another limitation is the potential for re-identification in white-box scenarios, where an attacker familiar with the pitch-shifting method could reverse-engineer the obfuscation. Exploring adaptive models to counteract such attacks could improve security.

In conclusion, our study highlights pitch-shifting as a useful method for mitigating privacy risks in acoustic feature analysis for cognitive assessment. By identifying optimal balancing points between privacy preservation and diagnostic utility, we have provided a foundation for future research into more sophisticated obfuscation methods for digital health applications. Our approach contributes to the ongoing discussion on privacy in medical data analysis and promotes ethical practices in the use of voice data for cognitive health assessment.

## Data Availability

Data from the Dementia Bank Delaware corpus can be downloaded from the public domain. Data from FHS (https://www.framinghamheartstudy.org/fhs-for-researchers/data-available-overview/) can be obtained by contacting and conditions for access include the successful completion of all steps outlined at https://www.framinghamheartstudy.org/fhs-for-researchers/, as well as approval from the FHS Research Committee.

## Acknowledgements

We gratefully acknowledge all participants of the Framingham Heart Study and Dementia Bank cohorts for their invaluable contributions to this research. We extend our thanks to the dedicated researchers and staff involved in the meticulous data collection, management, and analysis processes, whose collaborative efforts have been essential to advancing our understanding in this area. We also thank our institutional partners and funding agencies for their support, which has been fundamental in enabling this work.

## Consent statement

All participants included in this study provided informed consent, with the understanding that their speech data would be used for research purposes related to cognitive assessment. This study has been reviewed and approved by the relevant institutional ethics committee, confirming that it adheres to guidelines for responsible data handling and participant privacy.

## Figures and tables captions

**Supplementary Figure 1. Heatmap analysis of the privacy-cognitive tradeoff model applied to the FHS and Dementia Bank datasets**. (a) Correlation between pitch-shifted level, equal error rate, average classification accuracy, AUC speaker recognition, and total performance ρ(α=0.5) for the FHS dataset. (b) Correlation between pitch-shifted level, equal error rate, average classification accuracy, AUC speaker recognition, and total performance ρ(α=0.5) for the Dementia Bank dataset.

## References

1. Mahon E, Lachman ME. Voice biomarkers as indicators of cognitive changes in middle and later adulthood. Neurobiology of Aging. 2022;119:22–35. doi: 10.1016/j.neurobiolaging.2022.06.010.

2. Ginsberg SD, Themistocleous C, Eckerström M, Kokkinakis D. Voice quality and speech fluency distinguish individuals with Mild Cognitive Impairment from Healthy Controls. Plos One. 2020;15(7). doi: 10.1371/journal.pone.0236009.

3. Amini S, Hao B, Zhang L, Song M, Gupta A, Karjadi C, Kolachalama VB, Au R, Paschalidis IC. Automated detection of mild cognitive impairment and dementia from voice recordings: A natural language processing approach. Alzheimers Dement. 2022. Epub 20220707. doi: 10.1002/alz.12721. PubMed PMID: 35796399; PMCID: PMC10148688.

4. Chen J, Ye J, Tang F, Zhou J. Automatic Detection of Alzheimer’s Disease Using Spontaneous Speech Only. Interspeech 20212021. p. 3830–4.

5. Ding H, Lister A, Karjadi C, Au R, Lin H, Bischoff B, Hwang PH. Detection of Mild Cognitive Impairment From Non-Semantic, Acoustic Voice Features: The Framingham Heart Study. JMIR Aging. 2024;7. doi: 10.2196/55126.

6. Haider F, de la Fuente S, Luz S. An Assessment of Paralinguistic Acoustic Features for Detection of Alzheimer’s Dementia in Spontaneous Speech. IEEE Journal of Selected Topics in Signal Processing. 2020;14(2):272–81. doi: 10.1109/jstsp.2019.2955022.

7. Hajjar I, Okafor M, Choi JD, Moore E, Abrol A, Calhoun VD, Goldstein FC. Development of digital voice biomarkers and associations with cognition, cerebrospinal biomarkers, and neural representation in early Alzheimer’s disease. Alzheimer’s & Dementia: Diagnosis, Assessment & Disease Monitoring. 2023;15(1). doi: 10.1002/dad2.12393.

8. Karjadi C, Xue C, Cordella C, Kiran S, Paschalidis IC, Au R, Kolachalama VB. Fusion of Low-Level Descriptors of Digital Voice Recordings for Dementia Assessment. J Alzheimers Dis. 2023;96(2):507–14. doi: 10.3233/JAD-230560. PubMed PMID: 37840494; PMCID: PMC10657667.

9. Xue C, Karjadi C, Paschalidis IC, Au R, Kolachalama VB. Detection of dementia on voice recordings using deep learning: a Framingham Heart Study. Alzheimers Res Ther. 2021;13(1):146. Epub 20210831. doi: 10.1186/s13195-021-00888-3. PubMed PMID: 34465384; PMCID: PMC8409004.

10. Lin K, Washington PY. Multimodal deep learning for dementia classification using text and audio. Scientific Reports. 2024;14(1). doi: 10.1038/s41598-024-64438-1.

11. Nishikawa K, Akihiro K, Hirakawa R, Kawano H, Nakatoh Y. Machine learning model for discrimination of mild dementia patients using acoustic features. Cognitive Robotics. 2022;2:21–9. doi: 10.1016/j.cogr.2021.12.003.

12. Shi M, Cheung G, Shahamiri SR. Speech and language processing with deep learning for dementia diagnosis: A systematic review. Psychiatry Research. 2023;329. doi: 10.1016/j.psychres.2023.115538.

13. Yang Q, Li X, Ding X, Xu F, Ling Z. Deep learning-based speech analysis for Alzheimer’s disease detection: a literature review. Alzheimer’s Research & Therapy. 2022;14(1). doi: 10.1186/s13195-022-01131-3.

14. Javeed A, Dallora AL, Berglund JS, Ali A, Ali L, Anderberg P. Machine Learning for Dementia Prediction: A Systematic Review and Future Research Directions. Journal of Medical Systems. 2023;47(1). doi: 10.1007/s10916-023-01906-7.

15. Pulido MLB, Hernández JBA, Ballester MÁF, González CMT, Mekyska J, Smékal Z. Alzheimer’s disease and automatic speech analysis: A review. Expert Systems with Applications. 2020;150. doi: 10.1016/j.eswa.2020.113213.

16. Nautsch A, Jiménez A, Treiber A, Kolberg J, Jasserand C, Kindt E, Delgado H, Todisco M, Hmani MA, Mtibaa A, Abdelraheem MA, Abad A, Teixeira F, Matrouf D, Gomez-Barrero M, Petrovska-Delacrétaz D, Chollet G, Evans N, Schneider T, Bonastre J-F, Raj B, Trancoso I, Busch C. Preserving privacy in speaker and speech characterisation. Computer Speech & Language. 2019;58:441–80. doi: 10.1016/j.csl.2019.06.001.

17. Lanzi AM, Saylor AK, Fromm D, MacWhinney B, Cohen ML. Establishing the DementiaBank Delaware Corpus: An Online Multimedia Database for the Study of Language and Cognition in Dementia. Alzheimer’s & Dementia. 2023;19(S19). doi: 10.1002/alz.073058.

18. Lanzi AM, Saylor AK, Fromm D, Liu H, MacWhinney B, Cohen ML. DementiaBank: Theoretical Rationale, Protocol, and Illustrative Analyses. American Journal of Speech-Language Pathology. 2023;32(2):426–38. doi: 10.1044/2022_ajslp-22-00281.

19. Mahmood SS, Levy D, Vasan RS, Wang TJ. The Framingham Heart Study and the epidemiology of cardiovascular disease: a historical perspective. The Lancet. 2014;383(9921):999–1008. doi: 10.1016/s0140-6736(13)61752-3.

20. McFee B, Raffel C, Liang D, Ellis D, McVicar M, Battenberg E, Nieto O. librosa: Audio and Music Signal Analysis in Python. Proceedings of the 14th Python in Science Conference2015. p. 18–24.

21. Savitzky A, Golay MJE. Smoothing and Differentiation of Data by Simplified Least Squares Procedures. Analytical Chemistry. 2002;36(8):1627–39. doi: 10.1021/ac60214a047.

22. Jyh-Min C, Hsiao-Chuan W. A method of estimating the equal error rate for automatic speaker verification. SympoTIC ‘04 Joint 1st Workshop on Mobile Future & Symposium on Trends In Communications (IEEE Cat No04EX877)2004. p. 285–8.

