## Supplementary figures and images for "Obfuscation via pitch-shifting for balancing privacy and diagnostic utility in voice-based cognitive assessment"

### Supplemental figure 1

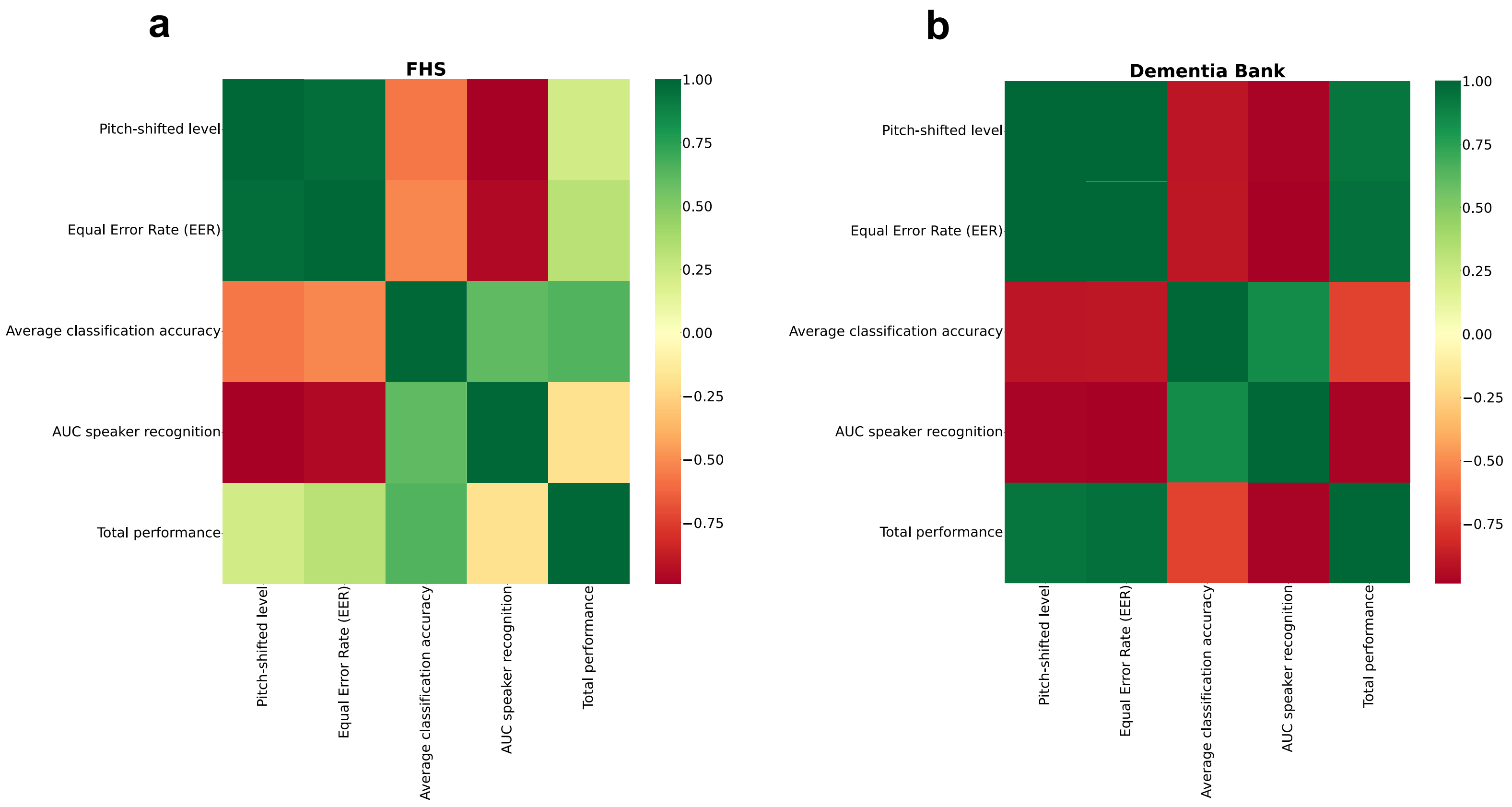
