## Supplementary tables for "Obfuscation via pitch-shifting for balancing privacy and diagnostic utility in voice-based cognitive assessment"

**Supplementary Table 1.** Classification accuracy of six algorithms using original and top-20 acoustic features, employing 10-fold cross-validation method on FHS and Dementia Bank datasets.

| **Dataset** | **Random forest** | **SVM** | **KNN** | **MLP** | **Ada boost** | **Gaussian**  **Naive Bayes** | **Average accuracy** |
| --- | --- | --- | --- | --- | --- | --- | --- |
| **FHS** | Original = 0.6705  Top-20 = 0.6846 | Original = 0.6929  Top-20 = 0.6929 | Original = 0.5994  Top-20 = 0.6622 | Original = 0.5936  Top-20 = 0.5359 | Original = 0.559  Top-20 = 0.5686 | Original = 0.4917  Top-20 = 0.5526 | Original = 0.6012  Top-20 = 0.6161 |
| **Dementia bank** | Original = 0.6194  Top-20 = 0.8 | Original = 0.6014  Top-20 = 0.6014 | Original = 0.5417  Top-20 = 0.7167 | Original = 0.4778  Top-20 = 0.7514 | Original = 0.5486  Top-20 = 0.722 | Original = 0.6736  Top-20 = 0.7847 | Original = 0.5771  Top-20 = 0.7294 |

**Supplementary Table 2.** Maximum total performance *ρ* of the tradeoff model across various privacy levels *α* (0.05 to 1) on the FHS dataset. The optimal tradeoff point is achieved at privacy level *α* = 0.35, corresponding to pitch-shifted level 0.4.

| **Privacy level (*α*)** | **EER** | **AUC speaker recognition** | **Average classification accuracy** | **Maximum performance** | **Maximum performance pitch** |
| --- | --- | --- | --- | --- | --- |
| 0.05 | 0.3342 | 0.7468 | 0.646467 | 0.630853 | 0.4 |
| 0.1 | 0.3342 | 0.7468 | 0.646467 | 0.61524 | 0.4 |
| 0.15 | 0.3342 | 0.7468 | 0.646467 | 0.599627 | 0.4 |
| 0.2 | 0.3342 | 0.7468 | 0.646467 | 0.584013 | 0.4 |
| 0.25 | 0.3342 | 0.7468 | 0.646467 | 0.5684 | 0.4 |
| 0.3 | 0.3342 | 0.7468 | 0.646467 | 0.552787 | 0.4 |
| 0.35 | 0.3342 | 0.7468 | 0.646467 | 0.537173 | 0.4 |
| 0.4 | 0.3476 | 0.7099 | 0.639083 | 0.52249 | 0.95 |
| 0.45 | 0.3476 | 0.7099 | 0.639083 | 0.507916 | 0.95 |
| 0.5 | 0.3476 | 0.7099 | 0.639083 | 0.493342 | 0.95 |
| 0.55 | 0.3476 | 0.7099 | 0.639083 | 0.478768 | 0.95 |
| 0.6 | 0.3476 | 0.7099 | 0.639083 | 0.464193 | 0.95 |
| 0.65 | 0.3476 | 0.7099 | 0.639083 | 0.449619 | 0.95 |
| 0.7 | 0.3476 | 0.7099 | 0.639083 | 0.435045 | 0.95 |
| 0.75 | 0.3476 | 0.7099 | 0.639083 | 0.420471 | 0.95 |
| 0.8 | 0.3476 | 0.7099 | 0.639083 | 0.405897 | 0.95 |
| 0.85 | 0.3476 | 0.7099 | 0.639083 | 0.391323 | 0.95 |
| 0.9 | 0.3476 | 0.7099 | 0.639083 | 0.376748 | 0.95 |
| 0.95 | 0.3495 | 0.707 | 0.607683 | 0.362409 | 1 |
| 1 | 0.3495 | 0.707 | 0.607683 | 0.3495 | 1 |

**Supplementary Table 3.** Maximum total performance *ρ* of the tradeoff model across various privacy levels *α* (0.05 to 1) on the Dementia Bank data. The optimal tradeoff point is achieved at privacy level *α* = 0.3, corresponding to pitch-shifted level 0.85.

| **Privacy level (*α*)** | **EER** | **AUC speaker recognition** | **Average classification accuracy** | **Maximum performance** | **Maximum performance pitch** |
| --- | --- | --- | --- | --- | --- |
| 0.05 | 0.0136 | 0.9898 | 0.721983 | 0.686564 | 0.2 |
| 0.1 | 0.0136 | 0.9898 | 0.721983 | 0.651145 | 0.2 |
| 0.15 | 0.0136 | 0.9898 | 0.721983 | 0.615726 | 0.2 |
| 0.2 | 0.0136 | 0.9898 | 0.721983 | 0.580307 | 0.2 |
| 0.25 | 0.0136 | 0.9898 | 0.721983 | 0.544888 | 0.2 |
| 0.3 | 0.2908 | 0.7886 | 0.620833 | 0.521823 | 0.85 |
| 0.35 | 0.2908 | 0.7886 | 0.620833 | 0.505322 | 0.85 |
| 0.4 | 0.3254 | 0.7455 | 0.5986 | 0.48932 | 1 |
| 0.45 | 0.3254 | 0.7455 | 0.5986 | 0.47566 | 1 |
| 0.5 | 0.3254 | 0.7455 | 0.5986 | 0.462 | 1 |
| 0.55 | 0.3254 | 0.7455 | 0.5986 | 0.44834 | 1 |
| 0.6 | 0.3254 | 0.7455 | 0.5986 | 0.43468 | 1 |
| 0.65 | 0.3254 | 0.7455 | 0.5986 | 0.42102 | 1 |
| 0.7 | 0.3254 | 0.7455 | 0.5986 | 0.40736 | 1 |
| 0.75 | 0.3254 | 0.7455 | 0.5986 | 0.3937 | 1 |
| 0.8 | 0.3254 | 0.7455 | 0.5986 | 0.38004 | 1 |
| 0.85 | 0.3254 | 0.7455 | 0.5986 | 0.36638 | 1 |
| 0.9 | 0.3254 | 0.7455 | 0.5986 | 0.35272 | 1 |
| 0.95 | 0.3254 | 0.7455 | 0.5986 | 0.33906 | 1 |
| 1 | 0.3254 | 0.7455 | 0.5986 | 0.3254 | 1 |
